# Understanding the asymmetric spread and case fatality rate (CFR) for COVID-19 among countries

**DOI:** 10.1101/2020.04.21.20073791

**Authors:** Eldhose Iype, Sadhya Gulati

## Abstract

The severe acute respiratory syndrome coronavirus 2 (SARS-CoV-2) infections are rising rapidly every day in the world, causing the disease COVID-19 with around 2 million people infected and more than 100,000 people died so far, in more than 200 countries. One of the baffling aspects of this pandemic is the asymmetric increase in cases and case fatality rate (CFR) among countries. We analyze the time series of the infection and fatality numbers and found two interesting aspects. Firstly, the rate of spread in a region is directly connected to the population density of the region where the virus is spreading. For example, the high rate of increase in cases in the United States of America (USA) is related to the high population density of New York City. This is shown by scaling the cumulative number of cases with a measure of the population density of the affected region in countries such as Italy, Spain, Germany, and the USA and we see that the curves are coinciding. Secondly, we analyzed the CFR number as a function of the number of days, since the first death, and we found that there are two clear categories among countries: one category with high CFR numbers (around 10%) and the other category with low CFR numbers (2% to 4%). When we analyzed the results, we see that countries with lower CFR numbers more or less tend to have implemented active control measures such as aggressive testing, tracking down possible infections, effective quarantine measures, etc. Moreover, we did not see any convincing correlation between mortality rates and the median age of the population.

## 1 Introduction

The COVID-19 pandemic has brought almost every country in the world to a standstill with partial or complete lockdown, restricting movements of people, shutting down businesses, world economy facing the possibility of a depression. Since the first reported infection of severe acute respiratory syndrome coronavirus 2 (SARS-CoV-2) [18] in a seafood market in the city of Wuhan, Hubei, China [13] in December 2019, the virus has now spread to 206 countries as of today (14^*th*^ April 2020) and infected 1,873,265 people with a global death toll of 118,854 [9]. The world health organization (WHO) has declared the disease a pandemic on 11th of March 2020, by then there were more than 118,000 cases in 114 countries with 4,291 deaths[1]. The pandemic is rampaging through almost every country in the world with the United States of America (USA) and a few European countries such as Italy, Spain, France, Germany and the United Kingdom(UK) being the most affected ones. All these countries have reported more cases as well as deaths than the country of origin (China) of the virus [9]. Lin et.al. [16] predicted that a large scale pandemic was unavoidable in Europe and the USA, and it would peak between April and July 2020. The rates at which the virus is spreading in some countries are much higher than those in China, with the USA being the highest (around 235,000 cases in just two months [12]). Although, a number of studies are reported to predict and analyse the spread of this virus, most of them are limited to individual countries such as Iran[6], China[15, 30, 28, 3], Italy [22, 24], the USA [20, 11], with only a few studies [8, 17, 3] analyzing multiple countries. Pedersen and Meneghini [23] have tried to correlate country-specific variations in the spread of COVID-19 with containment measures taken. However, many exceptions, such as France, Spain, and the UK, are found. In this work, we correlate the rate of spread among various countries to the population density of the affected region.

Another interesting aspect of this pandemic is the varying case fatality rate (CFR) among countries [12, 4]. Some countries such as Spain and Italy report a CFR > 10%, whereas countries such as China, the USA and South Korea reporting a CFR < 4% with Singapore being the lowest. Such variations were attributed to uncertainties in the number of reported cases (due to asymptomatic cases) in countries [12], possible delays in reporting cases [8], and demographic and socio-economic profile [21]. Baud et.al. [5] suggested that the real fatality rate may be calculated by taking the ratio of death on a given date to the number of confirmed cases 14 days (incubation period) earlier. In their study, the initial CFR was 5.6% for China and a high of 15.2% for the rest of the world. Although, their estimate converged to the WHO estimate (5.5- 5.9%), when considered over a longer period, there are disagreements to this approach [26]. In this work, we show that there are only two groups of countries (assuming that the number of deaths reported is correct for countries) based on the CFR value, and most countries can be categorized into either of the two.

## 2 Analysis methodology

In this analysis, we used the case distribution data, downloaded on 14^*th*^ April 2020, from the European Centre for Disease Prevention and Control (ECDC) website [9]. The country-wise daily reported cases and deaths were extracted. Since the starting dates for each country in the database is different, we have shifted the data to a specific starting point based on a minimum number of cases or deaths. A python script is used for the entire analysis.

## 3 Results and discussion

### 3.1 Initial rise in cases

The starting phase of the pandemic for a list of most affected countries (after 10 cases) is given in Figure 1. The trend appears to be similar for almost all countries except that the sharp rise in cases happens at various stages for different countries. For example, Turkey, Iran, Italy, and Spain show a quicker rise in cases, and countries such as Germany, the USA, and Singapore show a delayed rise in cases. One outstanding example is Singapore, which has not allowed the trend to rise as fast as the majority of countries. This is most likely due to the number of measures taken by Singapore in controlling the outbreak[19]. Another country which has managed to escape the outbreak is South Korea, as it can be seen in the figure, the rate of rising in cases is reduced significantly after 35 days since the first ten cases. This is also understandable from the extent of efforts taken by the South Korean government to contain the outbreak[25]. Eventhough there are many factors, such as late detection of the pandemic, early control measures, testing rates, and socio-economic factors, which may influence the varying start of the rise in cases, the trend in most countries beyond 2000-4000 cases appears to be similar. Although India is very early in the pandemic, we have included it in the figure because of the very high population densities in Indian cities, which we will prove to be the deciding factor in the rise in cases.

**Figure 1:**
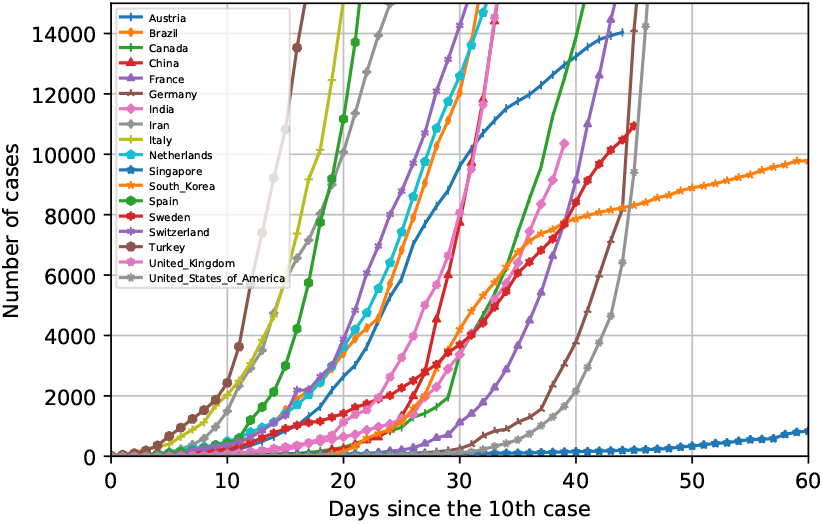
Cumulative number of cases for selected countries after 10 cases till 10000^*th*^ case. x-axis is the number of days since 10 cases are reported for each country.

### 3.2 Effect of population density on the spread of the virus (selected countries)

One of the surprising concerns in this pandemic is the varying rate of infection in different countries. Figure 2 a) shows the number of cases in selected countries (China, France, Germany, Iran, Italy, Spain, the UK, and the USA) after 2000 cases (we consider this as the growing phase of the pandemic). The cases in the USA are rising much faster than many other countries such as Italy, Spain, and Germany. One of the important factors which influences such a variation is the population density of the affected region. So we have adjusted the number of cases by population density as follows.

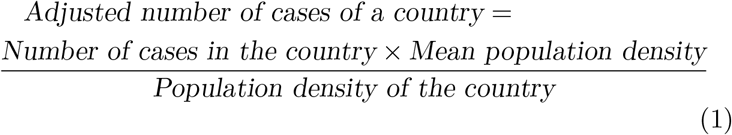

**Figure 2:**
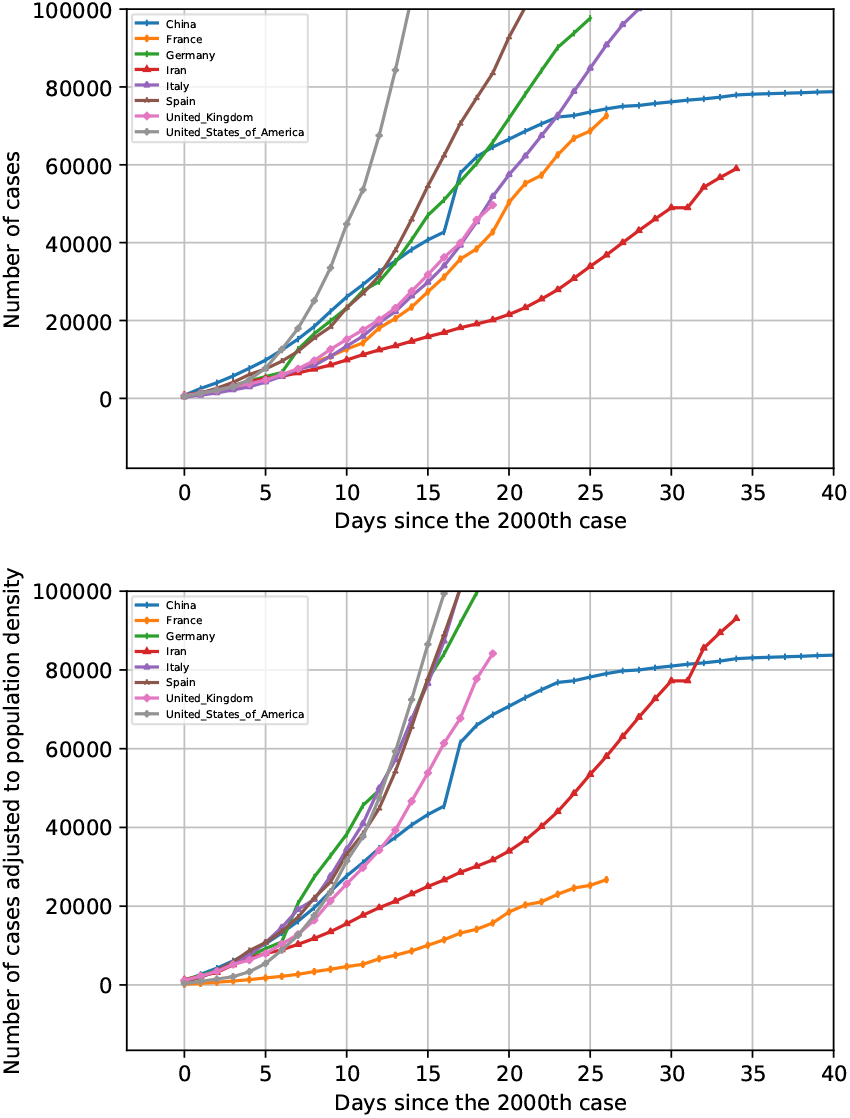
a) Cumulative number of cases after 2000 cases upto 100000 cases for selected countries. b) Number of cases adjusted for population densities of hot spots in each country.

However, instead of considering the population density of the country as a whole, we took the population density of the most affected region or city in those countries. So the cities that we chose and its population densities are given in Table 1

**Table 1:** Most affected cities in each country and their population densities

| Country | China | France | Germany | Iran | Italy | Spain | the UK | the USA |
| --- | --- | --- | --- | --- | --- | --- | --- | --- |
| Most affected city | Wuhan | Paris | Munich | Tehran | Bergamo | Madrid | London | New York |
| Population density (number of people per $km^2$) | 7242 | 20909 | 4668 | 4882 | 3000 | 5400 | 4542 | 10947 |

Figure 2 b) shows the adjusted number of cases based on population density for the selected countries and clearly, we see that at least four countries (Germany, the USA, Italy, and Spain) follow the same trend. Thus, the main reason for an accelerated rise in cases in the USA is the high population density of New York City (the most affected city in the USA). Besides, the initial part of China and the UK curves are almost following the same trend as the above four countries. However, China has managed to slow down the rise due to strict lockdown measures [14] and thus managed to flatten the curve. The trends in the UK, Iran, and France curves show that the rise in cases is smaller compared to other countries. Interestingly, the number of tests per million in these three countries (Iran:3421, France: 5114 and the UK: 5637)[2] are lower than the other selected countries (US: 8928, Italy: 17,315, Spain: 12,833, China: unknown)[2]. Nevertheless, this result shows that the rate of spread in a region is directly linked to population density and not to the overall population. Besides, the influence of crowded spots on the spread of the SARS-CoV-2 virus has been exemplified by the case of a church service in the city of Daegu [25], the main cause of the spread of the virus in South Korea. Since we could not pin point the hot spots in other countries and the unavailability of city-wise data, we did not extend this analysis for other countries. Moreover, this trend does not even hold for these four countries beyond 100,000 cases, because, by then the disease has spread to other parts within these countries. Perhaps a city-wise data of cases will probably help us understand more about this.

### 3.3 Case fatality rate (CFR) and two classes of countries

Next, we analyzed the case fatality rate (CFR) of COVID-19 for selected countries, which is one of the biggest unknowns of this pandemic. Some estimates can be found in the literature for various countries, however, our data suggest that there are only two clusters of countries based on the CFR data. We have calculated CFR as the ratio of the number of deaths to the number of confirmed cases on a given date. A plot of this as a function of days since the first death for a bunch of selected countries (the USA, the UK, Spain, Italy, China, Germany, France, and Iran) is shown in Figure 3 a). Firstly, we assume in this analysis that the number of deaths reported is equal or very close to the actual value, although this may not be true for many countries. Ideally, the CFR must be low in the beginning (because of incubation time and delay in developing complications from the infection) and then increases, and finds a plateau which will eventually become the ultimate CFR for the disease. From Figure 3 a), we observe two groups of countries emerging. One group with a high mortality rate (Spain, Italy, and the UK) and the other group with low mortality rate (the USA, Germany, China). The two countries which do not fall into these two are Iran and France. The starting point of this curve for Iran was at CFR=1 and for France, it is close to 0.09, which is equivalent to 100% and 9% mortality rates, respectively. A nine percent mortality is understandable, however, 100% is not. Thus, Iran must have started to report or test cases only after they saw fatalities due to the disease. Similarly, the initial part of these curves (localized peaks at third to fourth days from the 1st death) tells that contries such as France, China, the USA, and Italy reported relatively higher CFR values in the beginning of the pandemic. However, a notable exception is Germany, which started active testing of people with symptoms very early itself [27], which helped them identify cases faster than other countries and it managed to contain the eventual mortality rate to close to 2.5%. Because the curves for Iran and France are fluctuating significantly, we did not proceed with the data for these two countries. We saw in the previous section that the number of tests per million is also relatively low in these two countries.

**Figure 3:**
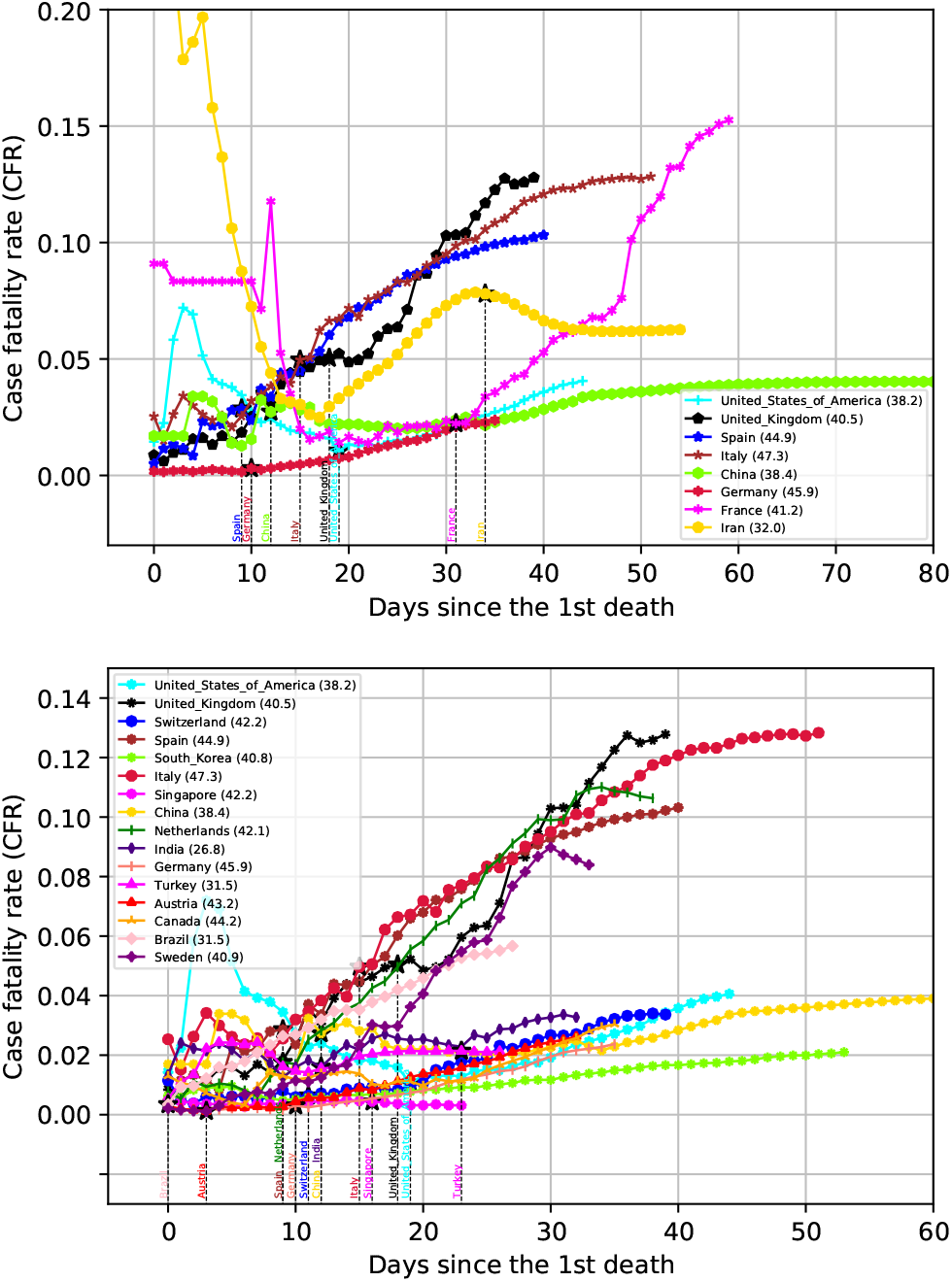
a) Case fatality rate (CFR) of selected countries. For Iran, the y-value starts at 1. b) CFR values of various countries show two clear trends. The numbers in the legend shows the median age of countries. The starting of containtment measures for countries are also indicated as vertical dotted lines.

Considering the high fluctations in the CFR for France and Iran (as discussed above), we have discarded those two countries and selected many other affected countries into our analyses and are shown in Figure 3 b). We see two trends among countries. One set of countries reports higher CFR values and settling at around 10% and the other group settling below 4%. We analyzed various factors that could explain such a split among countries. First of all, we looked at the demographics (median age of the countries are mentioned in the legend of Figure 3 b)). It can be argued that countries with more aged people may experience more deaths, however, we found as many exceptions (Brazil, Germany, Austria, Canada) as agreements (Spain, Italy, India, South Korea) to this argument. Another aspects we looked into is the geographic location and a possible influence of weather. However, some of these countries are geographically very close to each other (Germany vs. Netherlands, or Austria vs. Italy), yet the CFR curves are very different from each other. Next we analyzed the effect of containment measures on these curves. The dates of commencement of containment measures, if any, are marked marked for each country in Figure 3b). One interesting observation is that the countries which started the containment measures when the CFR values were low (Austria, Germany, Switzerland, Singapore, India, Turkey) have managed to contain the ultimate CFR, vs. countries which had higher CFR at lockdown (UK, Italy, Spain, Sweden (no lockdown)). These CFR values are shown in Table 2.

**Table 2:** CFR (%) at the date when containtment measures started.

| Country | CFR at lockdown (%) |
| --- | --- |
| Austria | 0.11 |
| Germany | 0.3 |
| Brazil | 0.34 |
| Singapore | 0.4 |
| Switzerland | 0.6 |
| the USA | 1.05 |
| India | 1.6 |
| Netherlands | 1.7 |
| Turkey | 2.1 |
| China | 2.7 |
| Spain | 2.8 |
| Italy | 4.9 |
| UK | 5.03 |
| Sweden | No lockdown |
| Canada | No lockdown |

For example, italy and the UK had CFRs of 4.9% and 5.03%, respectively, when the lockdown started. Both these countries are reporting high CFR currently. Some countries such as Germany[27], Singapore[19], and South Korea[25] were able to aggressively track, identify potential cases and quarantine them effectively. From Table 2, we can see that their CFR at lockdown was also low. Countries such as the USA and China[14, 10] implemented effective control methods which resulted in CFR number stabilizing at a decent level. There are exceptions to this rule as well. For example, the Netherlands and Brazil had early containment measures (as per wikipedia[7]) but reports higher ultimate CFR. For the case of Brazil, although the median age is lower compared to most of the countries, it appears that the possible difficulties in implementing effective lockdown of the country [20] may have contributed to the increasing number of deaths. Canada has less stringent containment measures, but effective control of CFR is probably due to the efficient healthcare system and adequate social distancing measures taken after the history with SARS epidemic [29]. Our analysis suggests that, surprisingly, a country like India with very high population densities in the cities, is reporting a CFR number below 0.4, which is heartening.

## 4 Conclusion

We have analyzed the COVID-19 cases and deaths of most affected countries using the data available on (14^*th*^ April) on the ECDC website and we have concluded the following.

- The variation in the rate of increase in cases in some countries such as the USA is related to the high population density of affected regions such as New York City. We see a direct correlation between the rate of spread with population density for at least four countries. Therefore, instead of analyzing overall population of a city or a country, one should consider the population density of the region or cities. This factor puts at risk some of the most populated cities in underdeveloped countries, urging them to take effective control action early enough to bring the pandemic under control.
- The case fatality rates were reported to be different for different countries in the literature. However, we have found two different trends in the CFR among various countries and most of the countries belong to either a higher ultimate CFR of close to 10% and the other group reporting an ultimate CFR value of between 2%-4%. We found no significant correlation between CFR and demographics (median age), but on the effective containment measures such as early detection of cases, tracking possible cases, effective quarantine measure, etc.

## Data Availability

The data used for this analysis was downloaded from ECDC website on 14th April 2020. This data is attached in the SI

https://www.ecdc.europa.eu/en/publications-data/download-todays-data-geographic-distribution-covid-19-cases-worldwide

